# Differential Metabolic Signatures of Cushing’s Disease Patients Dependent on their Obesity Status

**DOI:** 10.64898/2026.02.25.26346994

**Authors:** Treyton Carr, Irit Hochberg, Dave Bridges

## Abstract

Cushing’s disease is caused by the overproduction of cortisol. The effects of this disease are well known in a general population, including high blood pressure, diabetes, and weight gain. Cushing’s disease causes both obesity and metabolic related symptoms, and it can be difficult to discern the obesity-dependent from the obesity-independent mechanisms of Cushing’s disease. To identify patients with Cushing’s disease, we identified 476 Michigan Medicine patients between January 1^st^ 2000-2025 along with propensity-matched control cases.

We stratified our participants by obesity status and into a Cushing’s disease group and a control group. As expected, the Cushing’s group had an elevated BMI compared to the control group (34 kg/m^2^ vs 29 kg/m^2^). We found a higher proportion of females diagnosed with Cushing’s compared to males (287 vs 72). Cushing’s disease was associated with an increase in the fasting glucose levels in both non-obese and obese patients. In both the obese, and non-obese patients, there was an increase in ALT and AST levels regardless of Cushing’s disease status, but the increase due to Cushing’s disease was much greater in the patients with obesity (73.4 vs 35.1 mg/dL). Cushing’s disease also had a moderating effect on blood pressure, with participants a BMI under 30 kg/m^2^ increasing by 12.6 mmHg and participants with obesity increasing by only 7.9 mmHg. These findings highlight the need to consider obesity status when evaluating the effects of Cushing’s disease.

## Introduction

Cushing’s disease is a rare endocrine disorder, with an incidence rate of 1.8-2.5 cases per million people per year ^1,2^. This condition results from excess adrenocorticotropic (ACTH) hormone secretion from an adenoma in the pituitary gland, leading the adrenal glands to produce excess cortisol ^3^. Cushing’s disease is known to occur far more commonly in women, with an approximate female-to-male ratio of 4:1 ^1,4^. Patients with Cushing’s disease often experience hypertension, myocardial infractions, metabolic dysfunction associated steatotic liver disease (MASLD), weight gain, and hyperglycemia ^5–7^.

Weight gain, and obesity, are classic clinical manifestations of Cushing’s disease ^7,8^. Approximately 70-95% of patients experience weight gain, 21-48% of patients become overweight, and 32-41% of patients become obese ^5^. Development of obesity is also a leading cause of several of the Cushing’s related complications. As such, it is difficult to discern the obesity-independent from the obesity-dependent pathophysiology of Cushing’s disease. To address this gap, we performed a retrospective, case-control study from electronic medical records. We examined how obesity status modified the effects of Cushing’s disease by evaluating Cushing’s patients and matched controls with and without obesity.

## Materials and Methods

### Study Participants

This study was reviewed by the University of Michigan Institutional Review Board (HUM00247814) and adjudicated as not regulated. None of the investigators had access to identifiable patient data. To identify patients with Cushing’s disease, we evaluated electronic health records from participants at Michigan Medicine between January 1^st^ 2000-2025 searching for diagnoses of Cushing’s disease (including ICD Codes E24.0, and 255.0, validated in ^9^). We excluded participants if they did not have a Cushing’s disease diagnosis, treatment date, lab history, or relevant demographic data documented. Patients without a treatment date were excluded to ensure lab results were measured prior to and not after treatments. Race and ethnicity were self-reported, with groups of less than ten aggregated into the other reported categories listed in table 1.

**Table 1:**
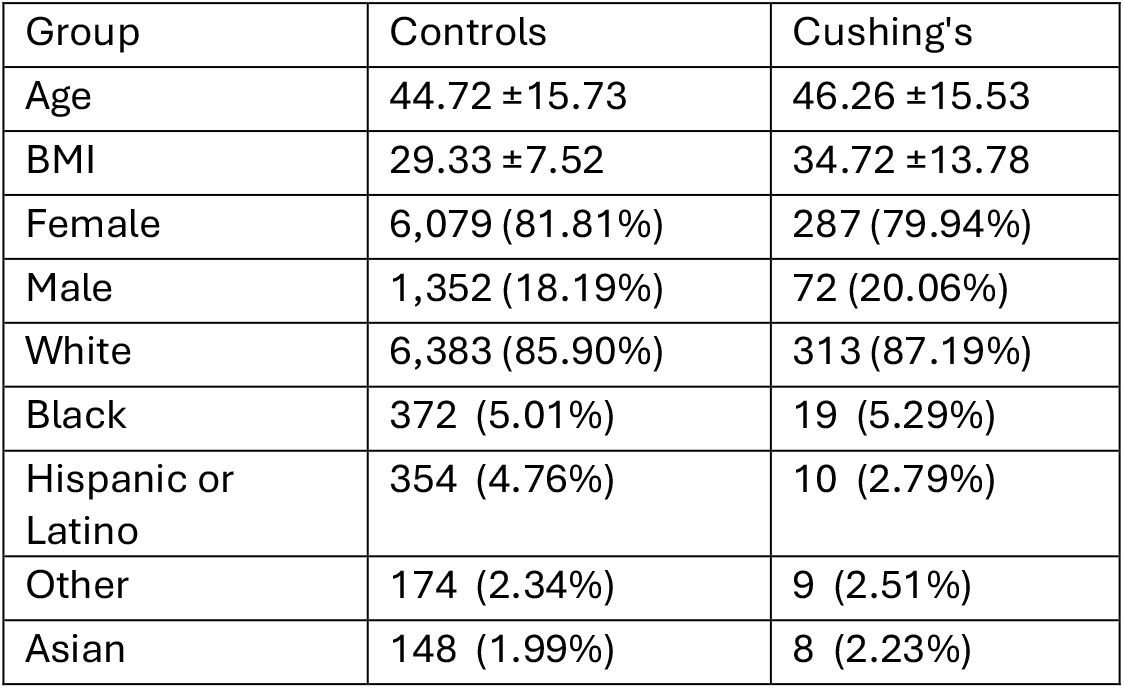
Participant characteristics after covariate matching. Participants were matched for age, gender and race/ethnicity but not BMI. This table represents the matched set for patients with Cushing’s disease with any laboratory value we considered. Supplementary Tables S2A-E describe outcome-specific matched characteristics.

| Group | Controls | Cushing’s |
| --- | --- | --- |
| Age | 44.72 ±15.73 | 46.26 ±15.53 |
| BMI | 29.33 ±7.52 | 34.72 ±13.78 |
| Female | 6,079 (81.81%) | 287 (79.94%) |
| Male | 1,352 (18.19%) | 72 (20.06%) |
| White | 6,383 (85.90%) | 313 (87.19%) |
| Black | 372 (5.01%) | 19 (5.29%) |
| Hispanic or Latino | 354 (4.76%) | 10 (2.79%) |
| Other | 174 (2.34%) | 9 (2.51%) |
| Asian | 148 (1.99%) | 8 (2.23%) |

To identify controls, we used the MatchIt package (version 4.7.1; ^10^). For each outcome we identified all Cushing’s cases with that measurement and potential controls with that measurement available. For Cushing’s cases to be included they had to have had the laboratory measured within a year prior to surgical treatment, taking the most recent value. We performed nearest‐neighbor propensity score matching (ratio = 10:1) with age in the propensity score model and exact matching on gender and race/ethnicity from a pool of 98,436 Michigan Medicine patients. This procedure was repeated separately for each outcome as our primary analyses. The maximal post-matching standardized mean difference for age was <0.05 for all outcomes, indicating adequate balance (Supplementary Figure 1 and Supplementary Table 1). We only included patients with BMIs within the 5-75 range, and between 18-75 years of age. Finally, we also only included non-Cushing’s patients as potential controls if they had at least one clinical visit in the 2022–2025-year range.

BMI was not included in the propensity score because we aimed to evaluate obesity as a moderator of the effects of Cushing’s disease. Subsequent analyses were stratified by obesity (BMI ≥30 kg/m^2^). Within each obesity stratum, BMI was included as a covariate in sensitivity models.

Mean arterial pressure (MAP) was directly reported in most cases. When MAP was missing but systolic and diastolic BP were available, MAP was calculated as (SBP + 2×DBP)/3. The calculated values showed high agreement with measured MAP (r=0.963, mean difference=1.32 ± 3.38 mmHg) in cases where both were available.

### Statistical Analyses

Statistical significance for this study was set at p<0.05. All analyses were performed using R version 4.4.0 ^11^. For participant demographics, Kruskal-Wallis or Mann-Whitney tests were used as both BMI and participant Age were not normally distributed (p<0.05 via Shapiro-Wilk tests). χ^2^ tests were used to compare counts across groups unless counts were expected to be under five per group, in which case Fisher’s exact test was used.

## Results

As described in Figure 1, we began with 5,428,302 participants with electronic health at Michigan Medicine. We found 2,893 unique diagnoses of Cushing’s disease. Next, we eliminated participants that did not have a documented treatment date after their diagnosis, giving us 476 participants. We then removed participants that did not have any of our tested laboratory results, giving us 453 participants. Finally, we removed a final set of participants that did not have complete demographic data, leaving us with our final participant population size of 365.

**Figure 1:**
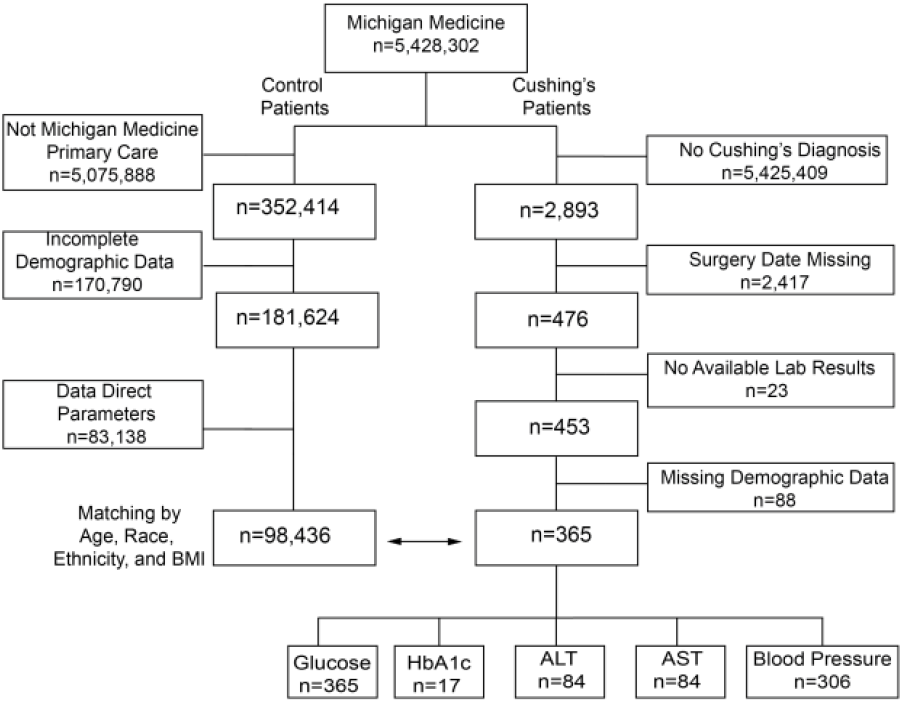
Participant flow diagram. Schematic showing the identification of Cushing’s and the pool of non-Cushing’s control patients used for each analysis.

Of these 365 participants, there had documented fasting glucose measurements for all 365, HbA1c values for 17, ALT values for 84, AST values for 84, and blood pressure data for 306 participants. The average age at diagnosis was 46.3 +/-15.5 years. There were 287 female and 72 male patients. The average BMI was 34.72 (+/-13.78). The patient population was predominately White (87.19%), with the remaining participants identifying as Black (5.29%), Hispanic or Latino (2.79%), Asian (2.23%), or Other (2.51%). These patients were then matched to controls without Cushing’s disease selected from a pool of 181,624 patients.

### Participant characteristics associated with Cushing’s disease

We performed outcome-specific propensity matching to identify appropriate controls, starting with generating exact matches for gender, race and ethnicity. We then algorithmically matched for age, generating ten controls for each case. Matching was effective with a standardized mean difference for age of <0.05 for each outcome (see Table 1, Supplementary Figure 1 and Supplementary Table S1).

Primary descriptive characteristics are provided for all participants with any available laboratory measurement (Table 1). Outcome-specific matched sets are described in Supplementary Tables S2A-E. There was a higher prevalence of Cushing’s disease in females (284 vs 72, p<0.001), consistent with the prior literature ^12–16^. As expected, we also observed a higher body mass index for patients with Cushing’s disease compared to controls (34.7 vs 29.3 kg/m^2^, p<0.001). Prior to matching for Race and Ethnicity, we noted that participants with Cushing’s disease were more likely to identify as Non-Hispanic White (87%) than the overall Michigan Medicine participant population (75%, p<0.001). Correspondingly, patients with Cushing’s disease were less likely to identify as Asian (p<0.001) or Black (p=0.018). This is consistent with other reports on the prevalence of Cushing’s disease across racial groups ^17,18^.

As we expected, participants with Cushing’s disease had a higher BMI than the controls, with 64% of participants with Cushing’s disease having a BMI above 30 kg/m^2^, compared to 40% controls (p<0.001). To understand the cross-sectional relationship between obesity and Cushing’s disease, we stratified our participants with a BMI above or below 30 kg/m^2^, across both cases and controls. We found that 232 patients had a BMI above 30 kg/m^2^ and 128 had a BMI below 30 kg/m^2^ at time of diagnosis. As shown in Table 2, and as expected given our stratification, the four groups differed in several characteristics, which were adjusted for in multivariable models below. For example, among the subgroup of participants with a BMI > 30 mg/m^2^, participants with Cushing’s disease had a higher BMI than controls (p=0.0024 by a Mann-Whitney test). Females with Cushing’s disease were more likely to have a BMI over 30 kg/m^2^ (68% of females compared to 51% of males with Cushing’s disease, p=0.015). Among participants with Cushing’s disease, Asian (6 out of 10) and Hispanic or Latino (6 out of 8) participants were more likely to have a BMI under 30 kg/m^2^ compared to the overall average (36%, p=0.027 and 0.176 respectively via Fisher’s exact tests). The average age of participants with Cushing’s disease with or without obesity was similar after stratification. Overall, the subgroup of patients with obesity and Cushing’s disease tended to be disproportionately female and White, but similar in age.

**Table 2:** Participant characteristics after stratification by obesity. Values are mean ± SD or n (%). Groups were stratified by Cushing’s status and obesity; no matching was performed across these strata. Kruskal-Wallis or χ^2^ tests compare across the four groups as appropriate.

| Variable | Control, BMI < 30 | Cushing's, BMI < 30 | Control, BMI > 30 | Cushing's, BMI > 30 | p-value |
| --- | --- | --- | --- | --- | --- |
| n | 4469 | 127 | 2962 | 232 |  |
| BMI | 24.47 ± 3.26 | 25.91 ± 3.16 | 36.67 ± 6.02 | 39.54 ± 14.92 | <0.0001 |
| Age | 44.04 ± 16.38 | 46.8 ± 17.27 | 45.74 ± 14.65 | 45.97 ± 14.52 | 0.00015 |
| Female | 3655 (81.8%) | 92 (72.4%) | 2424 (81.8%) | 195 (84.1%) | 0.041 |
| Male | 814 (18.2%) | 35 (27.6%) | 538 (18.2%) | 37 (15.9%) |  |
| White | 3850 (86.1%) | 105 (82.7%) | 2533 (85.5%) | 208 (89.7%) | <0.0001 |
| Black | 168 (3.8%) | 7 (5.5%) | 204 (6.9%) | 12 (5.2%) |  |
| Hispanic or Latino | 200 (4.5%) | 6 (4.7%) | 154 (5.2%) | 4 (1.7%) |  |
| Other | 120 (2.7%) | 3 (2.4%) | 54 (1.8%) | 6 (2.6%) |  |
| Asian | 131 (2.9%) | 6 (4.7%) | 17 (0.6%) | 2 (0.9%) |  |

### Modification of Cushing’s disease-related comorbidities by obesity

To identify how obesity alters metabolic outcomes related to Cushing’s disease, we evaluated the effects of Cushing’s disease on several key metabolic variables including glycemia (fasting glucose and for a smaller number of participants, HbA1c), liver enzymes, and blood pressure (Figure 2 and Table 3). As described below we noted substantial effect modification by obesity for liver enzymes and blood pressure (Table 3B).

**Table 3:** Effect sizes and statistical significance for each outcome. A) Sub-group specific effects of Cushing’s disease after adjusting for age, gender and race/ethnicity for each outcome. B) Summary of interactions between obesity and Cushing’s disease on each outcome, after adjusting for age, gender, and race/ethnicity. Data are presented as mean with 95% confidence intervals.

| A. Outcome | Obesity | Control | Cushing | Difference | p-value |
| --- | --- | --- | --- | --- | --- |
| Glucose | Non-Obese | 99.50<br>(98.31±100.70) | 124.17<br>(118.78±129.57) | 24.67<br>(19.32±30.02) | <0.0001 |
| Glucose | Obese | 107.67<br>(106.39±108.95) | 134.60<br>(130.48±138.73) | 26.93<br>(22.88±30.99) | <0.0001 |
| HbA1c | Non-Obese | 5.60<br>(5.24±5.95) | 6.51<br>(5.54±7.48) | 0.91<br>(-0.06±1.89) | 0.065 |
| HbA1c | Obese | 6.02<br>(5.68±6.35) | 7.40<br>(6.80±8.00) | 1.38<br>(0.82±1.95) | <0.0001 |
| ALT | Non-Obese | 23.40<br>(17.11±29.69) | 58.53<br>(45.27±71.79) | 35.13<br>(22.63±47.63) | <0.0001 |
| ALT | Obese | 29.21<br>(22.80±35.62) | 102.59<br>(91.24±113.95) | 73.39<br>(63.37±83.40) | <0.0001 |
| AST | Non-Obese | 23.00<br>(8.38±37.62) | 39.92<br>(9.06±70.77) | 16.91<br>(-12.16±45.99) | 0.25 |
| AST | Obese | 21.80<br>(6.90±36.69) | 78.73<br>(52.49±104.97) | 56.93<br>(33.85±80.02) | <0.0001 |
| Mean Arterial Pressure | Non-Obese | 85.73<br>(85.39±86.08) | 93.84<br>(91.51±96.17) | 8.11<br>(5.78±10.43) | <0.0001 |
| Mean Arterial Pressure | Obese | 90.04<br>(89.66±90.42) | 91.57<br>(89.79±93.34) | 1.53<br>(-0.24±3.29) | 0.090 |
| Systolic Blood Pressure | Non-Obese | 119.58<br>(119.07±120.09) | 131.55<br>(128.14±134.96) | 11.97<br>(8.57±15.36) | <0.0001 |
| Systolic Blood Pressure | Obese | 125.92<br>(125.37±126.47) | 129.00<br>(126.40±131.60) | 3.08<br>(0.50±5.66) | 0.019 |
| Diastolic Blood Pressure | Non-Obese | 68.78<br>(68.44±69.11) | 74.85<br>(72.58±77.11) | 6.07<br>(3.81±8.32) | <0.0001 |
| Diastolic Blood Pressure | Obese | 72.09<br>(71.72±72.45) | 72.67<br>(70.94±74.39) | 0.58<br>(-1.13±2.30) | 0.51 |

| B. Outcome | Interaction Estimate | p-value |
| --- | --- | --- |
| Glucose | 2.26 (-4.45±8.98) | 0.51 |
| HbA1c | 0.47 (-0.65±1.59) | 0.41 |
| ALT | 38.26 (22.26±54.26) | <0.0001 |
| AST | 40.02 (2.93±77.11) | 0.035 |
| Mean Arterial Pressure | -6.58 (-9.50±-3.66) | <0.0001 |
| Systolic Blood Pressure | -8.89 (-13.15±-4.63) | <0.0001 |
| Diastolic Blood Pressure | -5.48 (-8.32±-2.65) | 0.00015 |

**Figure 2:**
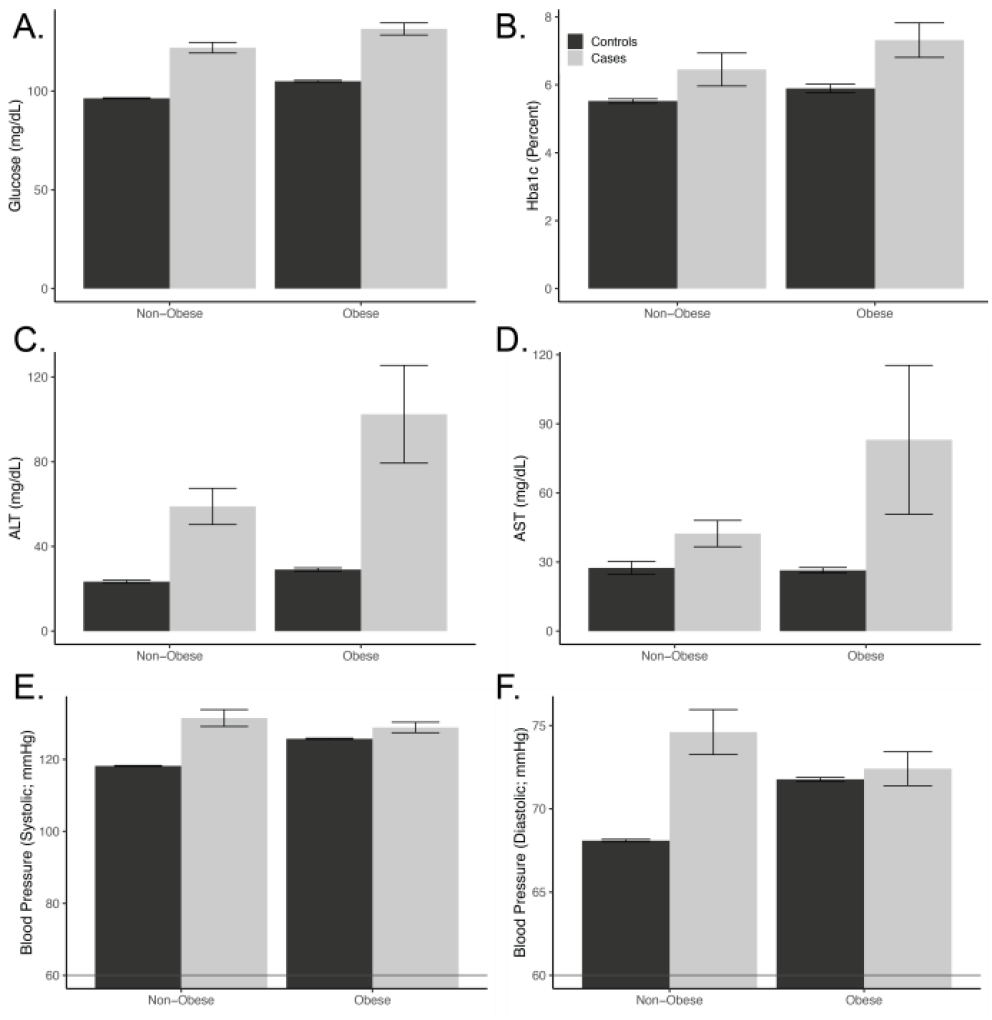
Effects of Cushing’s disease and Obesity on Laboratory Values. Laboratory values were within a year of Cushing’s disease surgery for cases. Obesity was defined as a BMI above or below 30 kg/m^2^. Data are presented as means with a 95% confidence interval, with lighter grey indicating data from participants with Cushing’s disease compared to propensity matched controls. Table 3 reports effect sizes and p-values for effects of Cushing’s disease and the interaction between disease and obesity.

#### Dysglycemia

The first metabolic variable of interest was the fasting glucose levels of our obese and non-obese populations. As shown in Figure 2 and Table 3A, we observed a significant increase in fasting blood glucose levels among participants with Cushing’s disease, regardless of obesity status (p<0.01 for both groups). When comparing glucose values of patients with BMI over 30 kg/m2 to those with a BMI under 30 kg/m^2^, Cushing’s disease patients and control patients had a similar glucose difference, suggesting that there is no effect modification of the excess glucocorticoids on the effect of obesity on glucose (p=0.51; Table 3B). Among the small number of patients with HbA1c values, there was an insignificant increase in HbA1c levels in the non-obese patients with Cushing’s disease compared to controls (0.91%; p=0.056). However, there was evidence of a significant increase in HbA1c levels in patients with obesity and Cushing’s disease compared to controls (1.38% p<0.01) (Table 3A). Again, there was no evidence of effect modification by obesity (p=0.41). This suggests that blood glucose increases with obesity and Cushing’s disease are largely additive.

#### MASLD and liver enzymes

To assess MASLD we looked at ALT and AST levels (Figure 2 and Table 3A). In both our obese and non-obese populations, we saw an increase in ALT and AST levels regardless of Cushing’s disease status (p<0.01 for both groups). In the case of ALT levels, the increase due to Cushing’s disease was much greater in the patients with obesity (73.4 vs 35.1 mg/dL, p_interaction_<0.01). This was also true for AST levels (increase by 56.3 mg/dL in patients with obesity and 16.9 for patients without obesity, p_interaction_=0.034). These data support the hypothesis that obesity enhances liver damage associated with Cushing’s disease.

#### Blood Pressure

We evaluated blood pressure readings as systolic, diastolic and mean arterial pressures (Figure 2 and Table 3A). We observed significant increases for all measures in patients with Cushing’s disease without obesity compared to controls without obesity (p=<0.01). We did not, however, observe this same significant magnitude of increases in blood pressure in Cushing’s patients with obesity population. For example, in terms of systolic blood pressure, Cushing’s disease was associated with an increase of 11.97 mm Hg in patients without obesity but 3.08 mm Hg in those with a BMI over 30 kg/m^2^. The attenuation of the effect of Cushing’s disease in patients’ obesity was significant for mean, systolic, and diastolic blood pressure (p_interaction_<0.01 for each). This suggests that obesity may have a protective effect on Cushing’s induced hypertension.

#### Sensitivity and secondary analyses

We performed a variety of sensitivity analyses to test the assumptions used in evaluating these effects, summarized in Table 4. By and large, these analyses show that the effects described above are robust to different analytical methods. For example, the synergistic effects of obesity and Cushing’s disease on liver enzymes was similar and significant across different demographic adjustments. We also found that propensity matching cases for BMI or including BMI as a covariate did not attenuate these effects. Within subgroups, we did notice some suggestive trends. For example, the synergistic (and antagonistic) effects of obesity were larger as obesity increased. For example, the interaction between Class I obesity and Cushing’s disease on ALT levels was -12.8 mg/dL but increased to +30.3 for Class II obesity and 80.7 for class III obesity. Since this analysis already accounted for a main effect of obesity status, this suggests that as obesity increases, the negative effect of Cushing’s disease on liver enzymes increased, while the protective effect on blood pressure was also trended to increase in magnitude. Stratifying by gender, we noticed that female patients generally had worse but more variable effect estimates. In the case of AST, the interaction between obesity and Cushing’s disease was estimated at an increase of 41.9 [-3.7-87.5] mg/dL beyond Cushing’s disease or obesity additively, but for males it was 25.8 [11.4-40.1] mg/dL.

**Table 4:**
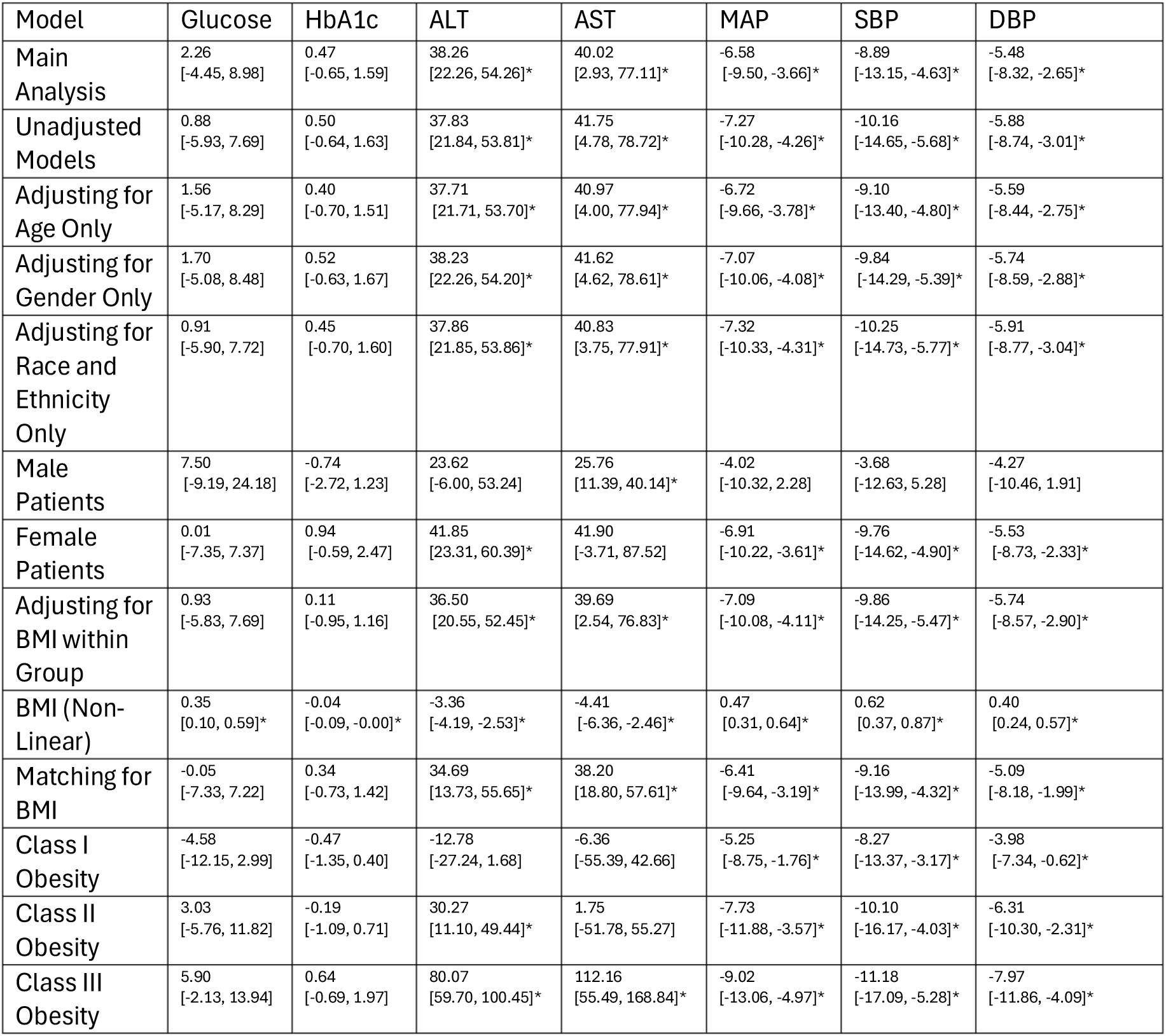
Sensitivity analyses for obesity-Cushing’s disease interactions. We report the estimate for the Cushing’s-Obesity interaction for each laboratory result as mean [95% confidence interval]. Each row describes a different analytical approach including subgroup analyses, and the inclusion of a more limited number of covariates. Unless otherwise noted all models adjusted for age, gender, and race/ethnicity. We also report several alternate models for obesity including adjusting for BMI within the primary stratified analyses, treating BMI as a non-linear covariate using non-linear splines, and including BMI as part of the propensity matching approach. Asterisks indicate p_interaction_<0.05.

## Discussion

This analysis of the effects of obesity and Cushing’s disease on metabolic complications demonstrates a complex set of interactions between the presence of obesity on Cushing’s disease- and obesity-related pathophysiology. We report that liver enzymes are enhanced relative to what we would predict from the additive effects of Cushing’s disease and obesity together, while in the case of blood pressure, obesity appears to reduce the effects of Cushing’s disease. Glucose homeostasis appears to be explained well by an additive model in our primary analysis where obesity and Cushing’s disease have non-synergistic effects. We see effects of both obesity and Cushing’s disease to increase blood glucose. Broadly, despite similar BMI levels between groups, metabolic dysfunction is dramatically worsened in patients with Cushing’s disease, so clearly there is substantial dysmetabolism beyond excessive weight gain.

These data are in part consistent with experimental animal studies in our group where pre-existing obesity aggravated liver steatosis and muscle function loss ^19,20^. These data are also consistent with our previous report demonstrating that obesity modifies Cushing’s-induced transcriptional changes in adipose tissue ^21^. We remain interested in Cushing’s induced bone and muscle loss, as well as the effect of obesity and Cushing’s disease on blood lipids and kidney function, but there are insufficient laboratory results in these participants to make reliable conclusions.

Pathophysiologically, the induction of adipocyte lipolysis by the direct effect of cortisol on adipocytes, as well as the indirect effects of cortisol on insulin signaling in adipocytes, may explain increased liver disease in patients with obesity and Cushing’s disease. This could be due to a confluence of increased triglyceride stores available for release, increased signaling, and decreased hepatic triglyceride export due to hepatic insulin resistance.

It is less clear to us why blood pressure is attenuated somewhat in patients with Cushing’s disease and obesity. Obesity and Cushing’s disease both increase systolic, diastolic and mean arterial pressure, but when both are combined, the effect is significantly attenuated (Figure 2 and Table 3). Obesity is a well-known risk factor for hypertension ^22–24^. Prior studies have shown 70-85% of Cushing’s disease patients suffer from hypertension in addition to the disease. One possibility is that patients with obesity and Cushing’s disease are already medicated to control their hypertension levels. Another possibility is there is already a ceiling effect as counter-regulatory mechanisms for blood pressure are already maximally engaged.

We also report several interesting gender differences in how obesity and Cushing’s disease interact. We find, as others have before, that Cushing’s disease affects women at a higher rate. We also find that women with Cushing’s disease are more likely to be obese. Women in this group, on average, tend to have worsened and more heterogeneous responses to these dual co-morbidities than males (Table 4). In our view, this warrants more detailed studies on sex differences, including the modifying effect of sex hormones on glucocorticoid signaling in the context of obesity. More broadly, even though obesity status was similar between the Cushing’s disease and control groups, patients with Cushing’s disease had more severe hyperglycemia, blood pressure and liver enzymes than those with just obesity.

There are several strengths of this study, including a large number of patients identified for a rare disease, coupled with standardized laboratory results and comprehensive medical records. The ability to match patients by age and demographic factors is also a strength of this analysis, as is the robustness of the analysis as demonstrated by our sensitivity analyses. In our primary analysis patients with Cushing’s disease and obesity tended to have a higher BMI than the non-Cushing’s patients with obesity (Table 2), raising a concern that perhaps the augmentation in ALT/AST was just due to patients being at a higher BMI. While this is a concern, we report that adjusting for BMI within group, and propensity matching for BMI did not modify the interaction effect estimates, which remained significant and approximately the same magnitude (Table 4), limiting that concern.

There are however several other limitations to this work. Many patients were diagnosed with Cushing’s disease, but their procedure date was not indicated. Second, this patient cohort was relatively homogeneous, consisting of mostly White patients, limiting our ability to evaluate race- or ancestry-specific differences. Underdiagnosis of nonwhite Cushing’s disease patients has been previously reported and remains plausible ^17^. Several patients did not have laboratory results in our window, so we were underpowered for some analyses (*i*.*e*. HbA1c) and unable to perform others (triglyceride levels, LDL levels, bone density) that were of interest. We appreciate that there also may be some surveillance bias in whom specific lab tests were reported, though these are relatively routine tests. In most cases, only one laboratory value was available prior to treatment, with the most recent value being used, but this precludes detailed time courses leading up to treatments. The long-term consequences of Cushing’s disease, with or without obesity are also highly important as patients may live decades after their treatment and we look forward to future studies to investigate these effects.

Cushing’s disease is unique in that it both causes obesity and causes several obesity-related pathophysiological comorbidities including MASLD, insulin resistance and hypertension. Our approach was geared towards separating the obesity- and Cushing’s dependent impacts on these co-morbidities and revealed a surprising and complex picture of integrated pathophysiology. This study provides novel data that obesity and Cushing’s disease interact, suggesting that obesity and Cushing’s disease may result in worsened liver damage, but that the augmented effects on hypertension are lower than expected. These results may be valuable for the accurate monitoring and treatment of the majority of Cushing’s patients who also have elevated adiposity.

## Supporting information

RECORD Checklist

## Data Availability

All code used for this analysis is available at https://github.com/BridgesLab/CushingAcromegalyStudy. Individual patient data cannot be shared for this study due to HIPAA privacy restrictions and ethical requirements.

## Acknowledgements and Funding

This work was supported by a pilot and feasibility grant from the Michigan Diabetes Research Center (NIH P30DK020572) and by consulting provided by the University of Michigan Adipose Tissue Core, part of the Michigan Nutrition and Obesity Research Center (NIH P30DK089503). This work was also supported by AI and Digital Health Innovation at the University of Michigan, and the University of Michigan Medical School Data Office for Clinical and Translational Research for providing data storage, management, processing, and distribution services. Finally, we would like to thank the other members of the Bridges and Hochberg laboratories for valuable discussions regarding these analyses and their interpretation.

