## Supplementary Figure Legends for "Differential Metabolic Signatures of Cushing’s Disease Patients Dependent on their Obesity Status"

### Supplementary Figure and Table Legends

**Supplementary Figure 1: Love plots showing covariate balance each outcome.** Plots describe the standardized mean difference for each variable prior to- (dark points) and after (light points) propensity matching. Dashed line indicates the standardized mean difference cutoff of 0.1. Note that age, race/ethnicity and gender were all exactly matched.

**Supplementary Table S1: Covariate matching diagnostics for age.** Averages, standard errors and standardized mean difference for age are shown for each outcome before and after propensity matching. Matching was exact for race and ethnicity.

**Supplementary Tables S2-6: Outcome-specific participant characteristics after matching**. Since propensity matching was completed for each Cushing’s patient with a laboratory value, the specific demographic characteristics for each analysis varies. For each analysis we report the demographics of those patients for [S2] Glucose, [S3] HbA1c, [S4] ALT, [S5] AST, and [S6] Blood Pressure.
