## Supplementary figures and images for "Differential Metabolic Signatures of Cushing’s Disease Patients Dependent on their Obesity Status"

### Supplementary Figures

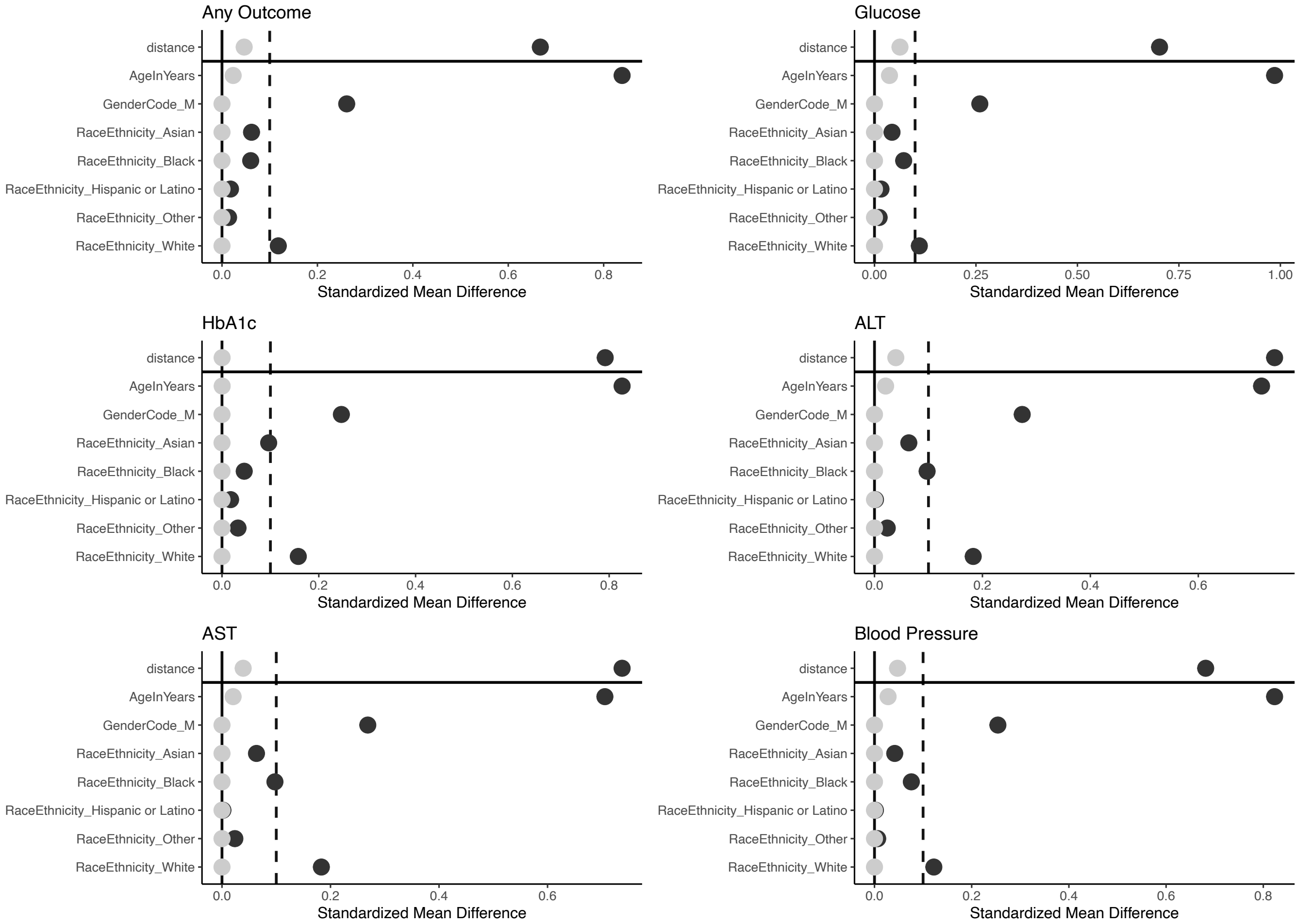
